# Retrospective cohort study of Ivermectin as a SARS-CoV-2 pre-exposure prophylaxis method in Healthcare Workers

**DOI:** 10.1101/2021.04.10.21255248

**Authors:** José Morgenstern, José N Redondo, Álvaro Olavarría, Isis Rondon, Santiago Roca, Albida De León, Juan Manuel Canela, Johnny Tavares, Miguelina Minaya, Óscar López, Ana Castillo, Ana María Plácido, Rafael Peña Cruz, Yudelka Merette, Marlenin Toribio, Juan Asmir Francisco

## Abstract

This observational and retrospective cohort study, carried out in two medical centers, Centro Medico Bournigal (CMBO) in Puerto Plata and Centro Medico Punta Cana (CMPC) in Punta Cana, Dominican Republic, sought to determine whether Ivermectin, at a weekly oral (PO) dose of 0.2 mg/kg, is an effective pre-exposure prophylaxis (PrEP) method preventing the spread of Severe Acute Respiratory Syndrome-Corona Virus-2 (SARS-CoV-2), in the healthcare workers. The study began on June 29, 2020 and ended on July 26, 2020, where 713 active healthcare personnel were included for the analysis, from which 326 healthcare personnel adhered to a weekly prophylactic program with Ivermectin for Coronavirus Disease-19 (COVID-19), designed by Grupo Rescue task force, that runs both medical centers, and 387 healthcare personnel who did not adhere to the program and were assigned as control group. A SPSS Propensity Score Matching procedure was applied in a 1:1 ratio to homogeneously evaluate the groups that were made up of 271 participants each.

In 28 days of follow-up, a significant protection of Ivermectin as a PrEP method preventing the infection from SARS-CoV-2 was observed 1.8% compared to those who did not take it 6.6%, (p-value = 0.006) with a risk reduction of a positive Chain Reaction of Real-Time Polymerase Transcription (RT-PCR) COVID-19 test by 74% (HR 0.26, 95% CI [0.10,0.71]). These results support the use of weekly Ivermectin as a compassionate preventive method in the healthcare personnel.

This study protocol is available at **Clinicaltrials**.**gov** Identifier: **NCT04832945**

## Introduction

Healthcare personnel are highly exposed to the SARS-CoV-2 infection. Therefore, it would be very wise and convenient to have a PrEP oral medication at hand, specially taking into consideration that worldwide mass vaccination against COVID-19, will probably not be achieved before the year 2022.

## Objectives

### Primary outcome measure

1. Number of participants with symptomatic SARS-CoV-2 infection and a positive COVID-19 RT-PCR test, in the Ivermectin group and in the control group.

### Secondary outcome measure

2. Number of sick participants with a RT-PCR COVID-19 positive test, whose condition deteriorated and required hospitalization and/or an Intensive Care Unit (ICU), in the Ivermectin group and in the control group.
3. Number of sick participants with a RT-PCR COVID-19 positive test who died, in the Ivermectin group and in the control group.

## Materials and methods

This study is an **observational retrospective multicenter cohort**, conducted on active **healthcare workers** at the **Centro Medico Bournigal** (CMBO) and the **Centro Medico Punta Cana** (CMPC), **Dominican Republic, started on June 29, 2020** and **ending on July 26, 2020**, followed up for 28 days, who were invited to a voluntary SARS-CoV-2 PrEP compassionate program using Ivermectin at a weekly PO dose of 0.2 mg/kg, that was orally administered at the Human Resources Office and were compared with a control group of healthcare personnel who did not receive Ivermectin, either by their own will or because contraindications in its use.

Both the Ivermectin group and the control group followed strictly the preventive measures and the use of the Personal Protection Equipment (PPE) to avoid the SARS-CoV-2 infection, implemented since April 2020, well before the start of these study. The SARS-CoV-2 symptomatic infected cases, were confirmed by a RT-PCR COVID-19 test.

The participants who adhered to the Ivermectin prophylaxis program had signed the informed consent, were minimum age of 18 years old, and were included in the intervention group only if: 1. received the first dose of Ivermectin during the first week of the study; 2. complied with a minimum of 2 doses of Ivermectin during the 4 weeks of the study; 3. the difference in days between the two doses was no greater than 14 days. The participants had weekly presentations in the Human Resources Office, where the medication was orally administered, being recorded the date of the Ivermectin delivery.

The following healthcare personnel were excluded from the Ivermectin prophylaxis program: pregnant or suspected pregnant women; women breastfeeding; patients receiving coumarin anticoagulants; those allergic to Ivermectin.

The healthcare personnel who did not adhere to the program until July 26, were assigned to the control group and at no time before and during the study did they take Ivermectin.

Those who had an RT-PCR COVID-19 positive, previous the start of the study, were excluded.

To obtain the demographic data of the workers, they were extracted from the Sigma software in CMPC and an internal development software in CMBO, from which the worker’s code, date of birth, sex and role or position were obtained. With this information, the Head of the Nursing Department assigned an exposure level, which was defined as follows:

1. High: personnel in direct contact with Covid-19 patients
2. Medium: personnel who care for non-Covid-19 patients
3. Low: personnel that does not take care of patients (administrative, others).

The compilation of the information from RT-PCR COVID-19 reports was done by consulting the Referencia Laboratorio Clínico system, located in Santo Domingo, Dominican Republic, who performed the analysis of the nasopharyngeal samples from both centers. Additionally, the healthcare personnel who gave an RT-PCR COVID-19 (+) during the study, were registered in their respective files in the Human Resources Office. The records of both medical centers, were consolidated into a single record by the Head of the Nursing Department, individualizing the worker, date of report and outcome (hospitalization and/or death).

Those participants in the study who tested positive for COVID-19, were treated as outpatients, and followed by their respective medical center with Ivermectin 0.4 mg/Kg PO one dose and Azithromycin 500 mg PO every 24 hours for 5 days. Those who required hospitalization received Ivermectin 0.3 mg/kg PO on days 1, 2, 6 and 7 plus Azithromycin 500 mg PO every 24 hours for 7 days. (1)

### Statistical analysis

To determine the degree of balance of the cohorts and to adjust the possible confounders of the study, a chi-square homogeneity test was performed. Those variables that obtained a p-value < 0.1 test were included in the Propensity Score Matching process of the SPSS software, in which a 1:1 match was performed with a tolerance of 0.05 to obtain homogeneous cohorts. These variables were gender, exposure and role.

After performing the matching, a chi-square test and a Kaplan-Meier hazard curve were performed with the positive COVID-19 test results of the healthcare personnel. Thereafter a Cox regression was performed to compare the effect of the study variables on the primary outcome which was a positive COVID-19 test. The validation and creation of the cohorts was done in MariaDB 10. The statistical analysis was done with IBM SPSS and RStudio 4.0.4

## Results

Initially 943 candidates were selected for the study, from which 43 were excluded for having a positive COVID-19 RT-PCR report before the start of the study. The Ivermectin group was made up of 510 healthcare personnel and the control group was made up of 390 participants. In total, 182 healthcare workers who did not comply with the Ivermectin doses described in the inclusion criteria were excluded from the Ivermectin group. In the follow-up study, 5 participants were lost, who resigned or were dismissed from their work, 2 in the Ivermectin group and 3 in the control group. This equates to a 0.6% loss of the follow-up for the Ivermectin group and 0.77% for the control group. As it is less than 5%, it is not considered relevant for the purposes of the study.

Finally, 713 participants were chosen, 326 in the Ivermectin group and 387 in the control group were considered for the preliminary analysis of this study. After the matching process with the Propensity Score, there were 271 members in each group for analysis (Figure 1).

**Figure 1:**
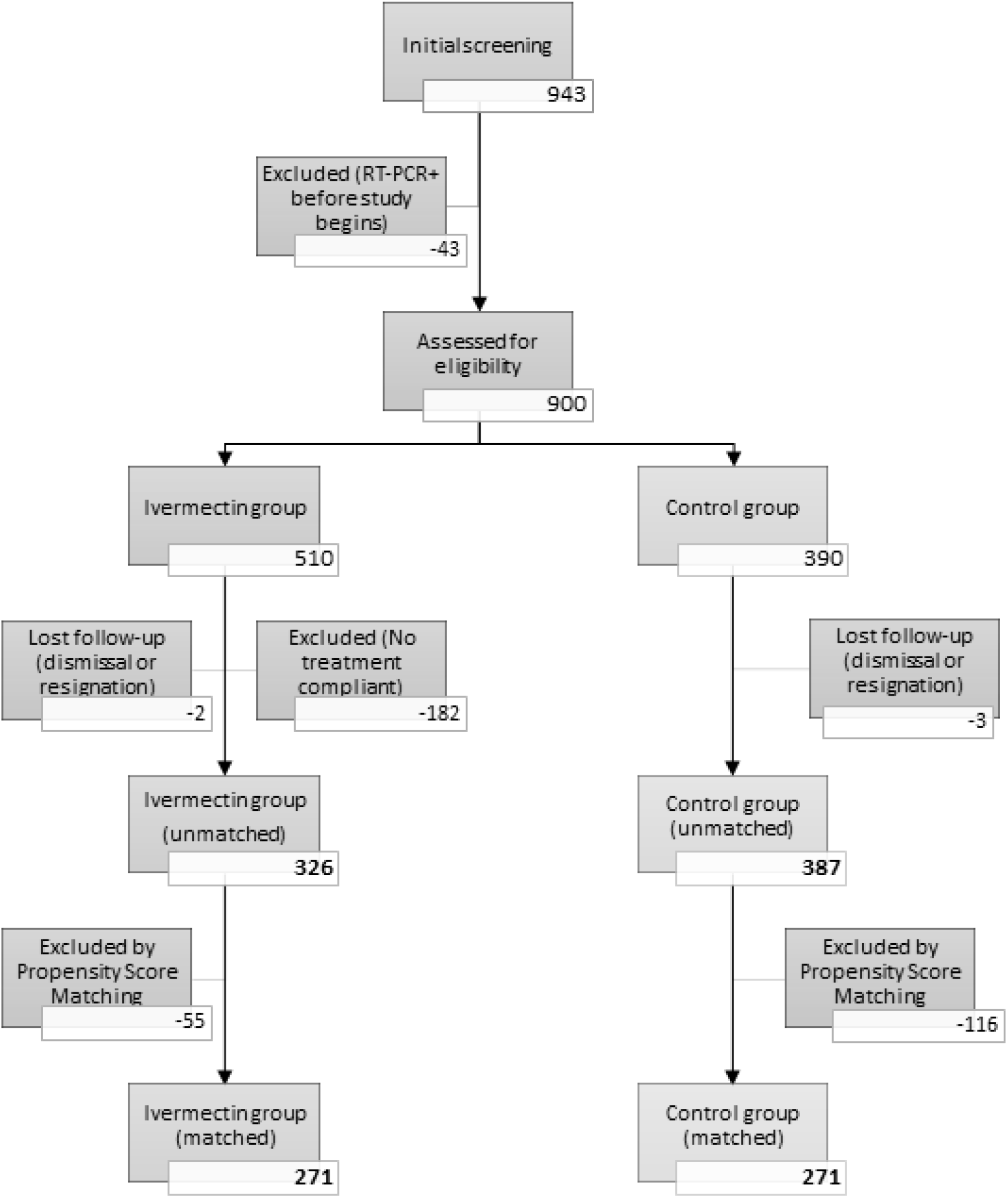
Evolution of the cohorts throughout the study flowchart.

The 51.5% of the participants in the Ivermectin group took 4 doses; 40.4% took 3 doses; 6.7% took 2 doses; 1.4% of the participants managed to take a single dose and then reported a positive COVID-19 infection (Figure 2).

**Figure 2:**
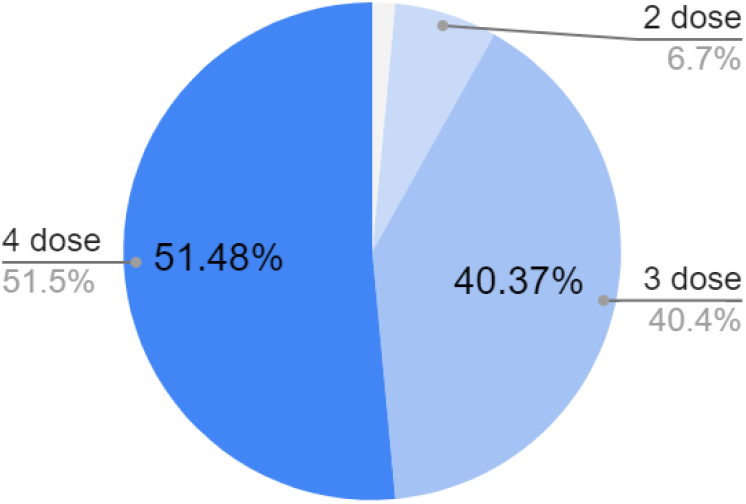
Percentage of the number of doses received by study participants.

There were no severe side effects reported from the use of Ivermectin. Only minor side effects such as dizziness (3.7%), headache (1.5%), stomachache (1.4%), pruritus (1.1%), nausea (1.1%) and diarrhea (0.7%).

The median time between doses was 7 days and the mean (average) was 7.2 days with a standard deviation of 1.03 (Figure 3).

**Figure 3:**
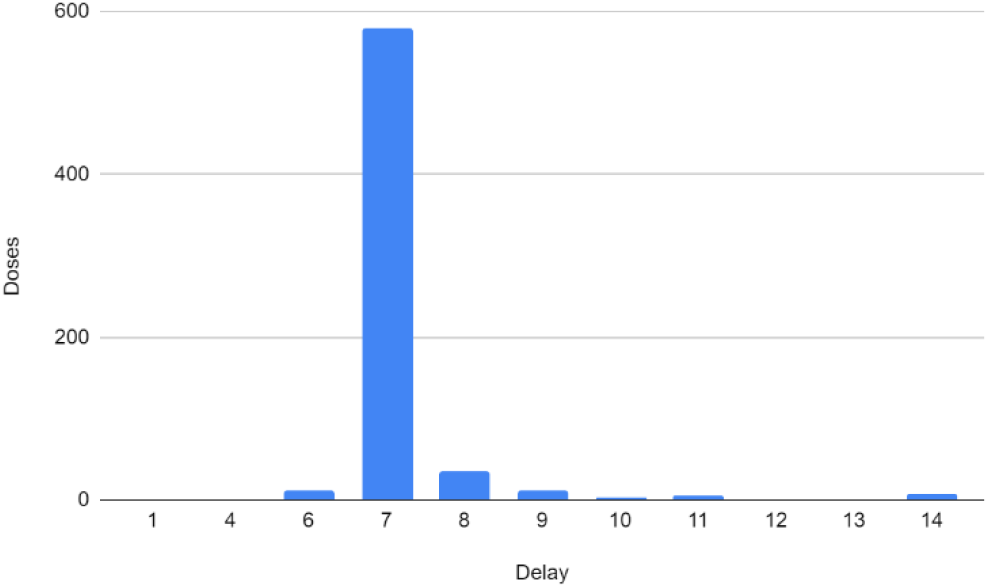
Time distribution between each dose (days).

### Demographic data

Executing the Propensity Score Matching process using the gender, exposure and role variables, the groups were left without significant differences in the measured variables (Table 1).

**Table 1:**
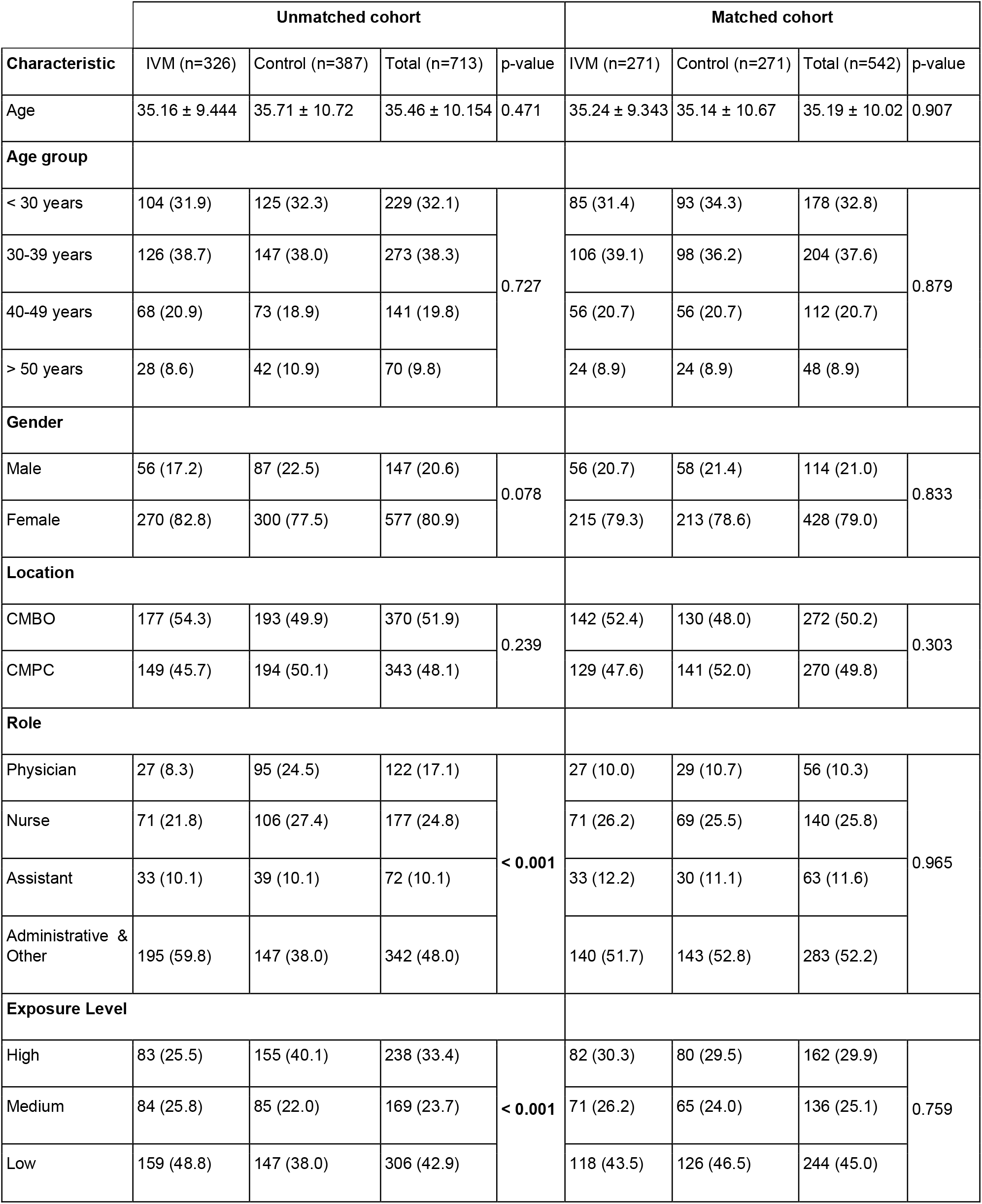
Chi-square homogeneity test of demographic characteristics of the participants at the beginning of the study. Unmatched and matched groups comparison.

|  | Unmatched cohort |  |  |  | Matched cohort |  |  |  |
| --- | --- | --- | --- | --- | --- | --- | --- | --- |
| Characteristic | IVM (n=326) | Control (n=387) | Total (n=713) | p-value | IVM (n=271) | Control (n=271) | Total (n=542) | p-value |
| Age | 35.16 ± 9.444 | 35.71 ± 10.72 | 35.46 ± 10.154 | 0.471 | 35.24 ± 9.343 | 35.14 ± 10.67 | 35.19 ± 10.02 | 0.907 |
| Age group |  |  |  |  |  |  |  |  |
| < 30 years | 104 (31.9) | 125 (32.3) | 229 (32.1) | 0.727 | 85 (31.4) | 93 (34.3) | 178 (32.8) | 0.879 |
| 30-39 years | 126 (38.7) | 147 (38.0) | 273 (38.3) |  | 106 (39.1) | 98 (36.2) | 204 (37.6) |  |
| 40-49 years | 68 (20.9) | 73 (18.9) | 141 (19.8) |  | 56 (20.7) | 56 (20.7) | 112 (20.7) |  |
| > 50 years | 28 (8.6) | 42 (10.9) | 70 (9.8) |  | 24 (8.9) | 24 (8.9) | 48 (8.9) |  |
| Gender |  |  |  |  |  |  |  |  |
| Male | 56 (17.2) | 87 (22.5) | 147 (20.6) | 0.078 | 56 (20.7) | 58 (21.4) | 114 (21.0) | 0.833 |
| Female | 270 (82.8) | 300 (77.5) | 577 (80.9) |  | 215 (79.3) | 213 (78.6) | 428 (79.0) |  |
| Location |  |  |  |  |  |  |  |  |
| CMBO | 177 (54.3) | 193 (49.9) | 370 (51.9) | 0.239 | 142 (52.4) | 130 (48.0) | 272 (50.2) | 0.303 |
| CMPC | 149 (45.7) | 194 (50.1) | 343 (48.1) |  | 129 (47.6) | 141 (52.0) | 270 (49.8) |  |
| Role |  |  |  |  |  |  |  |  |
| Physician | 27 (8.3) | 95 (24.5) | 122 (17.1) | < 0.001 | 27 (10.0) | 29 (10.7) | 56 (10.3) | 0.965 |
| Nurse | 71 (21.8) | 106 (27.4) | 177 (24.8) |  | 71 (26.2) | 69 (25.5) | 140 (25.8) |  |
| Assistant | 33 (10.1) | 39 (10.1) | 72 (10.1) |  | 33 (12.2) | 30 (11.1) | 63 (11.6) |  |
| Administrative & Other | 195 (59.8) | 147 (38.0) | 342 (48.0) |  | 140 (51.7) | 143 (52.8) | 283 (52.2) |  |
| Exposure Level |  |  |  |  |  |  |  |  |
| High | 83 (25.5) | 155 (40.1) | 238 (33.4) | < 0.001 | 82 (30.3) | 80 (29.5) | 162 (29.9) | 0.759 |
| Medium | 84 (25.8) | 85 (22.0) | 169 (23.7) |  | 71 (26.2) | 65 (24.0) | 136 (25.1) |  |
| Low | 159 (48.8) | 147 (38.0) | 306 (42.9) |  | 118 (43.5) | 126 (46.5) | 244 (45.0) |  |

The mean age of the study was 35.19 years (±10.02), comprised mainly of females (79.0%). Regarding the profession or role, physicians represent 10.3%, nurses 25.8%, medical assistants 11.6% and administrative personnel and others 52.2%. More than half of the personnel had medium and high exposure to COVID-19, corresponding to 29.9% and 25.1% (Table 1).

Comorbidities were not included in the demographic data as the information was not complete in 8.1%. However, with the existing data, this variable did nor represent an imbalance between the Ivermectin group and the control group (chi-square homogeneity test p-value=0.890) and in the Cox regression it also did not show to be a variable that affects the main outcome (p-value= 0.851, HR=1.1, 95% CI [0.4, 2.9]).

### Primary and secondary outcomes

The chi-square homogeneity test showed that PrEP with Ivermectin was associated with a statistically significant reduction in SARS-CoV-2 infection, with 1.8% versus 6.6% in the control group (p-value = 0.006) (Table 2).

**Table 2:**
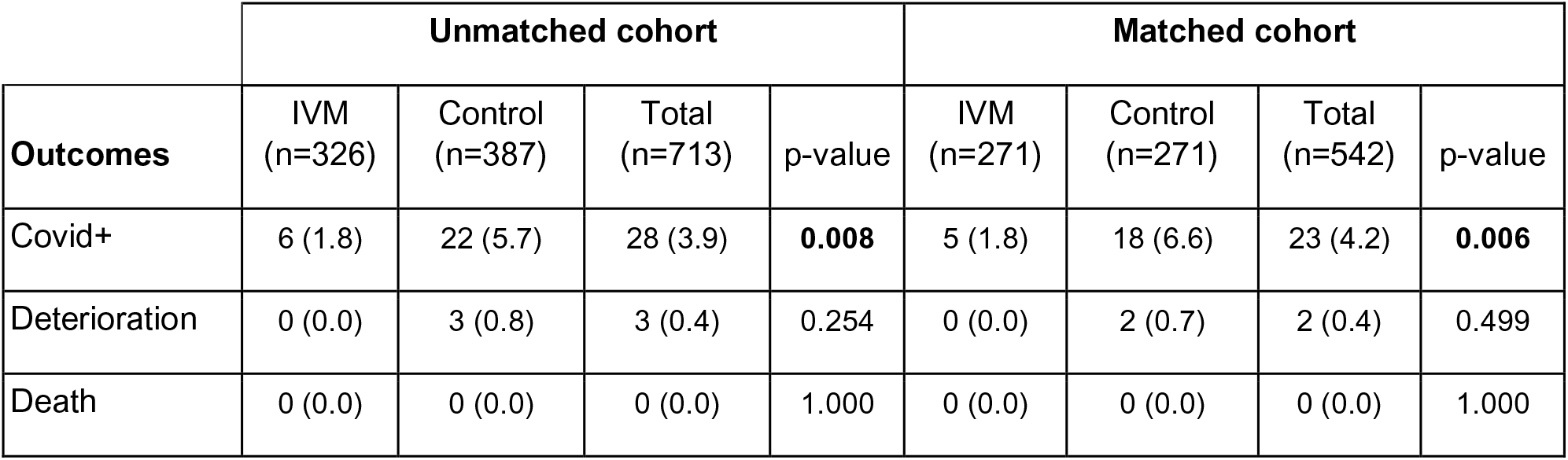
Chi-square homogeneity test outcomes 95% CI. Comparison of unmatched and matched groups. Fisher exact test for deterioration outcome p-value.

|  | Unmatched cohort |  |  |  | Matched cohort |  |  |  |
| --- | --- | --- | --- | --- | --- | --- | --- | --- |
| Outcomes | IVM<br>(n=326) | Control<br>(n=387) | Total<br>(n=713) | p-value | IVM<br>(n=271) | Control<br>(n=271) | Total<br>(n=542) | p-value |
| Covid+ | 6 (1.8) | 22 (5.7) | 28 (3.9) | **0.008** | 5 (1.8) | 18 (6.6) | 23 (4.2) | **0.006** |
| Deterioration | 0 (0.0) | 3 (0.8) | 3 (0.4) | 0.254 | 0 (0.0) | 2 (0.7) | 2 (0.4) | 0.499 |
| Death | 0 (0.0) | 0 (0.0) | 0 (0.0) | 1.000 | 0 (0.0) | 0 (0.0) | 0 (0.0) | 1.000 |

No progression of Covid-19 disease or complications which deserved hospital admission were observed and there were no mortalities, in the Ivermectin group. In Control Group a 0.7% showed clinical complications that warranted hospitalization, but no death. When comparing both groups the difference was not statistically significant in deterioration that required hospitalization and or ICU (p-value =0.499) and in death (p-value =1.000) (Table 2).

The Cox regression analysis showed that Ivermectin reduced the risk of COVID-19 infection by 74% compared to the control group (HR 0.26, 95% Cl [0.10, 0.71]). None of the analyzed covariables, like age, sex, workplace location (CMBO or CMPC), occupation, degree of exposure to COVID-19, reduced or increased significantly the risk of developing a COVID-19 infection (Table 3).

**Table 3:** Multivariate analysis (Cox regression) on COVID-19 infection outcome, 95% CI. Unmatched and matched groups comparison.

|  | Unmatched Cohort |  |  |  | Matched Cohort |  |  |  |
| --- | --- | --- | --- | --- | --- | --- | --- | --- |
|  | 95% CI for HR |  |  |  | 95% CI for HR |  |  |  |
| Variables | Hazard ratio | Lower | Upper | p-value | Hazard ratio | Lower | Upper | p-value |
| Age | 1.00 | 0.97 | 1.04 | 0.882 | 0.99 | 0.94 | 1.03 | 0.551 |
| Gender (Male) | 1.63 | 0.65 | 4.10 | 0.300 | 1.12 | 0.37 | 3.36 | 0.838 |
| Location (CMBO) | 1.61 | 0.75 | 3.49 | 0.224 | 1.38 | 0.59 | 3.23 | 0.455 |
| Role (Physician) | 0.74 | 0.18 | 3.07 | 0.681 | 1.22 | 0.26 | 5.67 | 0.802 |
| Role (Nurse) | 1.27 | 0.39 | 4.18 | 0.692 | 0.93 | 0.26 | 3.38 | 0.909 |
| Role (Assistant) | 0.44 | 0.07 | 2.75 | 0.380 | 0.42 | 0.07 | 2.69 | 0.362 |
| Exposure (high) | 2.32 | 0.59 | 9.12 | 0.228 | 3.81 | 0.82 | 17.64 | 0.088 |
| Exposure (medium) | 2.49 | 0.72 | 8.56 | 0.148 | 3.19 | 0.82 | 12.36 | 0.094 |
| Group (Ivermectin) | 0.33 | 0.13 | 0.83 | **0.019** | 0.26 | 0.10 | 0.71 | **0.008** |

Kaplan-Meier risk analysis showed that at 7^th^ day there were 7 patients with a COVID-19 infection in the control group and 3 patients in the Ivermectin group; during the second week there were 6 patients with a COVID-19 infection in the control group and 1 patient in the Ivermectin group. On third week there were 4 patients with a COVID-19 infection in the control group and 1 patient in the Ivermectin group. Finally, on fourth week there were 1 patient with a COVID-19 infection in the control group and 0 patient in the Ivermectin group (Figure 4).

**Figure 4:**
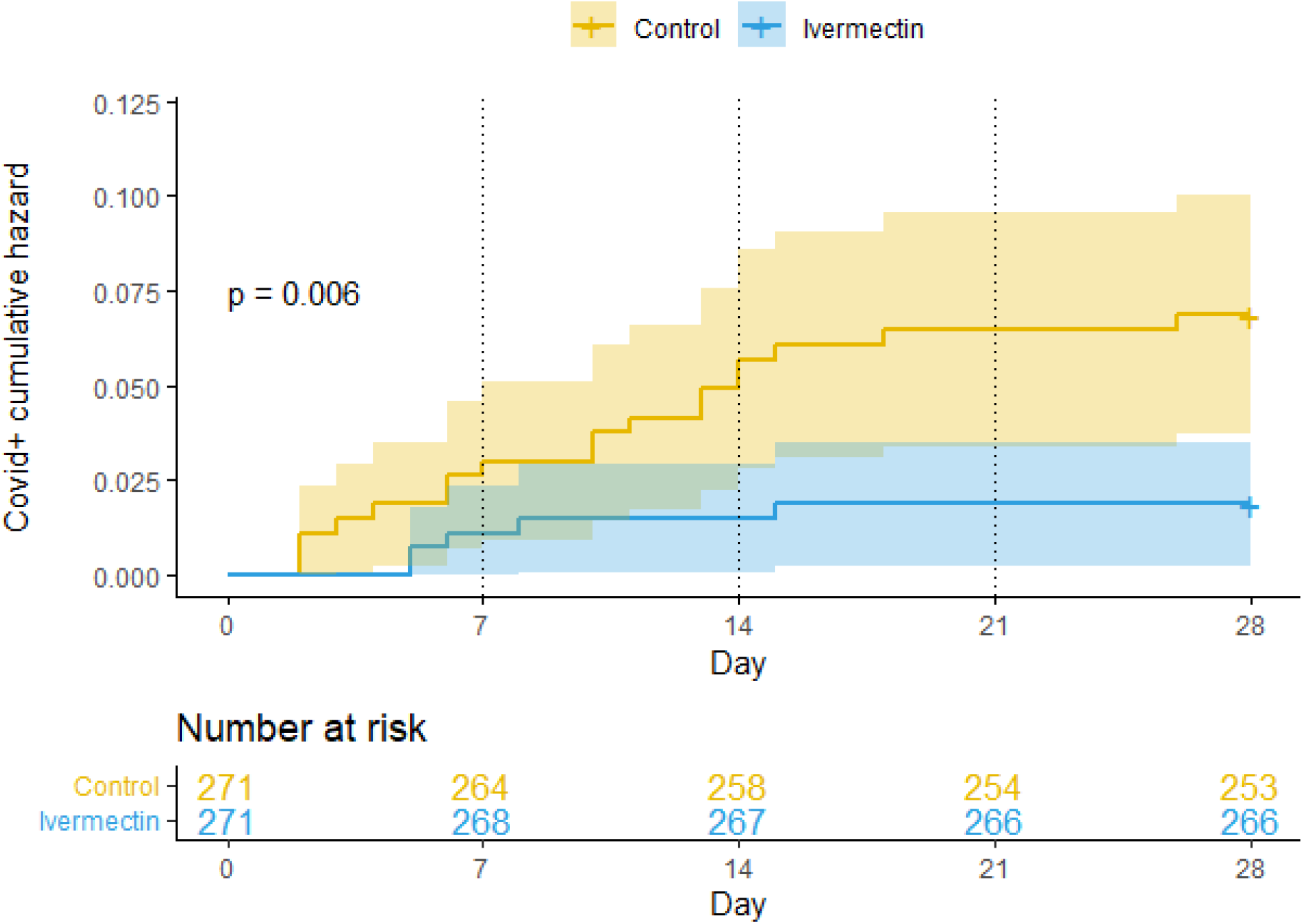
Kaplan-Meier cumulative risk curves for getting COVID-19 infection over the 28 days of this study. Log-rank method for p-value (95% CI).

The first 4 days after starting the study, a slightly higher number of positive COVID-19 infections was seen in the control group compared to the Ivermectin group. Then, from the fifth day, the number of positive COVID-19 infections is the same in both groups. At day 10, a difference between the two groups begins to be noticed, which increases over the course of the days until reaching day 28, where the protection of Ivermectin achieves significant statistics (p-value = 0.006). Between days 16 and 28 (12 days) the Ivermectin group did not present any contagion (Figure 4).

## Discussion

Ivermectin has 19 hours plasmatic half-life; an enterohepatic cycle and its active metabolites can remain in the blood for up to 72 hours. Its excretion is 99% hepatic and only 1% is excreted in the urine. If we give successive doses of Ivermectin on days 1, 4 and 7, the drug accumulation in blood is minimal (2, 3). Ivermectin is very lipophilic and its concentration in some tissues are high; active levels in the lungs reach almost 3 times higher than in the blood and is detectable in fat tissue for at least 7 days (4).

Kaplan-Meier risk analysis showed that the prevention with Ivermectin occurred after the second dose was received on day 8. This finding could suggest that to achieve a preventive dose of Ivermectin in the tissues, a second dose is needed (Figure 4).

This is consistent with the study results conducted in India, by Behera and colleagues, which evaluated 91 healthcare workers exposed to patients with COVID-19, who received preventive Ivermectin at 0.3 mg/kg on days 1 and 4, and were followed for one month. They compared this group with another group of 67 health workers, that was not administered Ivermectin. A 73% decrease risk in COVID-19 infection was observed in the Ivermectin group versus the placebo group (OR = 0.26, 95% CI [0.14, 0.47]). COVID-19 infections in the group that received the 2 doses of Ivermectin were not severe in any case and there were no cases of death. They reported a third group of 17 people who only took Ivermectin on day 1, in whom the preventive effect was not observed (5).

Chahla and colleagues, in Tucuman Argentina, conducted a 1:1 randomized controlled study in healthcare workers for 4 weeks with a 2-week follow-up after completion of the study. They compared a group that was administered PrEP with Ivermectin PO 12mg weekly plus Iota-Carrageenan 6 nasal sprays daily, versus a control group which did not receive any medication. In the “Ivermectin-Carrageenan” group, 4 out of the 117 participants (3.4%) were infected with COVID-19, and in the control group 25 out of the 117 participants (21.4%) were infected. The difference between the two groups was statistically significant (p-value <0.05). The odd ratio (OR) was 0.13, this value transformed into relative risk (RR) becomes 84% less risk of contagion of SARS-CoV-2 in the “Ivermectin-Carrageenan” group in comparison with the control group. In the Ivermectin group 4 patients had a mild infection and in the control group, in 15 patients the infection was mild, in 7 moderate and in 3 severe (6).

In our study, the hazard ratio (HR) of 0.26 was achieved in the Ivermectin group, which translates into a 74% lower risk of contagion with SARS-CoV-2. This risk result is similar to Behera et al, but 10 points less that Chahla et al. study. This additional difference could be attributed to the use of Iota-Carrageenan.

In the present study, when comparing healthcare personnel who presented COVID-19 infection, the progression of the disease, the appearance of complications, the need for hospitalization and the occurrence of death, the difference between both groups was not statistically significant. These findings could be due to their average young age of 35 years old, where patients tend to have fewer complications with COVID-19 infections and that the patients of both groups, were offered early treatment with Ivermectin, which diminishes the progression of SARS-CoV-2 infection, as seen in previous studies (1).

Since this is not a randomized study, it does not allow us to clear certain confounding factors, such as the fact that the Ivermectin group could be made up of healthcare personnel more concerned with prevention in general, including greater personal protection measures and more careful use of the PPE, reducing the risks of contracting SARS-CoV-2. To analyze this argument, we must follow up the same participants of the Ivermectin group (Figure 5), who passed from the weekly dose to a monthly dose PO 0.4 mg/kg and were followed for 56 days (8 weeks). During these 56 days, most of the healthcare workers discontinued progressively the Ivermectin on their own will, for nonspecific reason (Figure 6). When the Ivermectin group was compared to the control group, there was no statistically significant difference, regarding new cases of COVID-19 infections. This finding makes unlikely the former argument.

**Figure 5:**
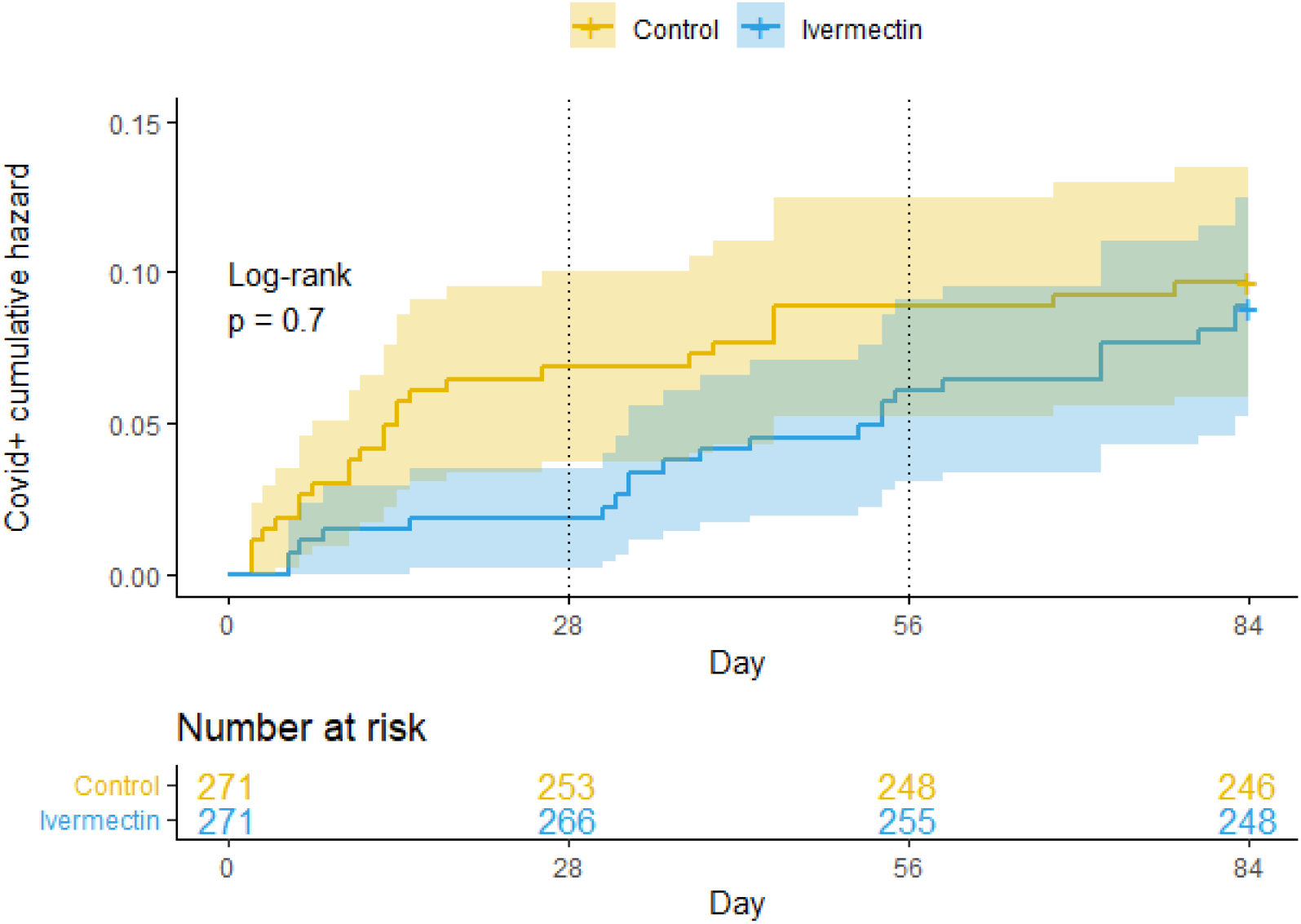
Kaplan-Meier cumulative risk curves for getting COVID-19 infection, extended version of Figure 4. The original 4 weeks of Ivermectin with a weekly dose and thereafter administering Ivermectin a monthly dose, during 8 weeks.

**Figure 6:**
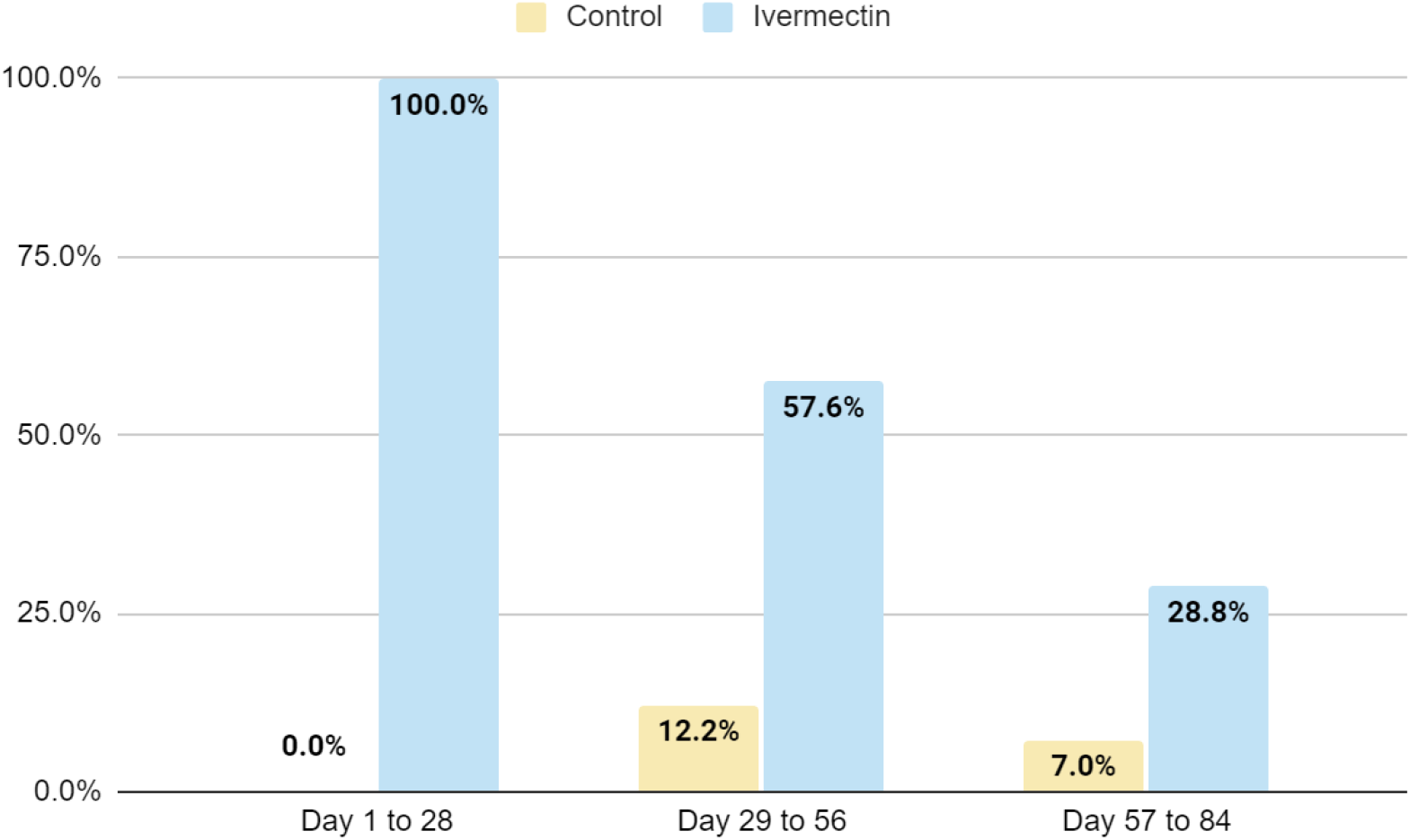
Ivermectin prophylactic program compliance for matched groups. The original 4 weeks with a weekly dose of Ivermectin, followed by 8 weeks with a monthly dose of Ivermectin.

Another element that may raise doubts could be the possibility that some participants in the control group used Ivermectin on their own or another preventive drug. We consider that this probability is very low, since the hospital management widely provided the medicine to the health personnel of both medical centers and free of charge.

## Limitations

1. The study was limited to 28 days, as in August 2020 the prevention scheme changed to a monthly dose of Ivermectin 0.4 mg/kg.
2. There was no RT-PCR test at the exit of this study, neither in the Ivermectin group nor in the control group. There is the possibility that asymptomatic cases were not detected.

## Conclusion

Pre-exposure prophylaxis (PrEP) for COVID-19, with a weekly oral dose of Ivermectin 0.2 mg/kg, was statistically significant in the exposed healthcare personnel, at the CMBO and CMPC, after 28 days of follow up, with only 1.8 % of the physicians and health collaborators developing SARS-CoV-2 infection versus 6.6% in the control group (p-value = 0.006). Ivermectin reduced the risk of contagion with COVID-19 by 74% compared to the control group (HR 0.26, 95% CI [0.10, 0.71]).

## Suggestions

1. The compassionate use of PrEP with Ivermectin 0.2 mg/kg PO per dose weekly, in countries where vaccination has not yet been possible should be considered, for the healthcare personnel, highly exposed to the SARS-CoV-2 infection.
2. Pre-exposure prophylactic double-blind, controlled randomized studies with different protocols, in workers highly exposed to the SARS-CoV-2 infection, comparing Ivermectin with other possible preventive treatments, would be desirable.

## Data Availability

The demographic data of the workers, they were extracted from the Sigma software in Centro Medico Punta Cana and an internal development software in Centro Medico Bournigal.
The compilation of the information from RT-PCR COVID-19 reports was done by consulting the Referencia Laboratorio Clinico system. The records of both medical centers, were consolidated into a single record by the Head of the Nursing Department, individualizing the worker, date of report and outcome (hospitalization and/or death).

## Acknowledgment

The authors wish to acknowledge all healthcare personnel of the participating institutions, for their invaluable professional dedication and interest in data collection.

